# Alignment and expansion in national adaptation of the WHO Model List of Essential Medicines: a cross-national analysis

**DOI:** 10.64898/2026.11.19.26368472

**Authors:** Yukari Kohno, Janne Estill, Lorenzo Moja, Olivia Keiser

## Abstract

**Background:** The WHO Model List of Essential Medicines (EML) provides a common global reference for national medicines selection. National adaptation frequently involves both the adoption of WHO-recommended medicines and the addition of medicines beyond WHO EML, which have not been distinguished in comparative analyses. This study quantifies how countries adapt the WHO EML by distinguishing alignment from expansion, exploring country-level characteristics potentially associated with these dimensions.

**Methods:** We compared national essential medicines lists (NEMLs) from 137 countries with the WHO EML. We constructed two independent measures: the Alignment Ratio, representing the proportion of WHO-recommended medicines included in each NEML, and the Expansion Ratio, representing the proportion of NEML medicines not included in the WHO EML. Countries were also classified into four adaptation modes based on combinations of alignment and expansion. Associations with country-level characteristics were examined using multivariable beta regression.

**Results:** Countries displayed substantial heterogeneity in adaptation patterns, with representation across all four adaptation modes defined by alignment with and expansion beyond the WHO EML. Fifty countries were categorized as restrictive/constrained adoption (low alignment, low expansion), thirty-five countries as normative dependence (high alignment, low expansion), sixteen countries as autonomous divergence (low alignment, high expansion) and thirty-six countries as capacity-driven expansion (high alignment, high expansion). Higher under-five mortality was associated with lower alignment (β = −0.294; 95% CI: −0.497 to −0.091) and lower expansion (β = −0.481; 95% CI: −0.697 to −0.264). Higher globalization index scores and more recent NEMLs were associated with higher alignment, but not expansion.

**Conclusion:** While some countries closely followed the WHO EML with limited additions, a substantial proportion combined high alignment with extensive expansion beyond the WHO benchmark, suggesting that national adaptation often involves supplementation rather than substitution of WHO recommendations.

## Background

The WHO Model List of Essential Medicines (EML) is a global normative instrument intended to support countries in selecting priority medicines and improving equitable access to essential treatments (Wirtz et al., 2017). First introduced in 1977, the EML has evolved from an experience-based list into an evidence-informed guidance instrument updated every two years through a WHO-coordinated review process (Laing et al., 2003).

Many countries have developed National Essential Medicines Lists (NEMLs) based on the WHO EML(Piggott et al., 2022). Over time the role of the Model List has evolved beyond its original focus on essential generic medicines in resource-constrained settings to include increasingly complex and high-cost therapies, prompting renewed debate regarding its purpose and implementation across diverse health systems (Jenei, 2025). Countries vary substantially in how closely their NEMLs align with the WHO EML and how extensively they add medicines beyond it. Such variation is expected because national medicine selection must reflect differences in disease burden, affordability, prescribing practices and health-system capacity(Persaud et al., 2019).

However, little is known about how national adaptation can be characterized quantitatively. Qualitative evidence has demonstrated that adaptation of the WHO EML involves complex processes of governance, evidence appraisal, financing, stakeholder engagement, and contextualization of global recommendations (Peacocke et al., 2022). Quantitative comparisons, by contrast, have largely treated differences between NEMLs and the WHO EML as a single measure. Previous studies found substantial variation in these differences and identified geographical region and health expenditure as important explanatory factors (Heredia et al., 2025; Persaud et al., 2019). However, this overall measure does not distinguish between the non-adoption of WHO-recommended medicines and the addition of medicines beyond the WHO baseline, potentially obscuring important differences in national medicines policy.

We therefore examine national adaptation through two independent but complementary dimensions: alignment with WHO-recommended medicines and expansion beyond the WHO benchmark. This distinction provides a potential framework for understanding how countries adopt and adapt the WHO EML across different health-system contexts.

## Methods

### NEMLs and WHO EML

We used a cross-sectional design to examine how countries adapt the WHO EML by comparing the contents of NEMLs against the WHO EML as a common global reference. The analysis was conducted at the medicine level, using the 2017 WHO EML, which contained 414 individual medicine entries(World Health Organization, 2017) , as the benchmark against which national lists were assessed. This reference period was selected because it enabled comparison with a large, geographically diverse, and systematically harmonized dataset of NEMLs. NEML data were derived from the Global Essential Medicines Database (Persaud et al., 2019), which contains medicine-level information from 137 NEMLs spanning all six WHO regions. Medicines were standardized using International Nonproprietary Names (INNs) and mapped to the Anatomical Therapeutic Chemical (ATC) classification system Level 4 (chemical subgroup) codes to facilitate cross-national comparison (World Health Organization, 2026).

### Outcome Measures

Consistent with our conceptualization of national adaptation as comprising two independent dimensions, we quantified adaptation to the WHO EML using measures of both alignment and expansion. Alignment captures the extent to which countries adopt medicines recommended by WHO, whereas expansion captures the extent to which countries include medicines beyond the WHO benchmark. As countries may simultaneously exhibit high levels of alignment and expansion, these measures were analysed separately.

For each NEML, medicines were classified as either aligned with the WHO EML or additional to it. Aligned medicines were those that matched a medicine included in the WHO EML, whereas additional medicines were those included in the NEML but not in the WHO EML.

The Alignment Ratio was calculated as the proportion of the 414 medicines included in the 2017 WHO EML that were also present in a country’s NEML. This measure reflects the degree to which a national list incorporates WHO-recommended medicines.

The Expansion Ratio was calculated as the proportion of medicines in a country’s NEML that were not included in the WHO EML. This measure reflects the extent to which national lists extend beyond the WHO benchmark.

Together, the two measures characterize national adaptation as a combination of adoption of WHO recommendations and addition of medicines reflecting national priorities, rather than as a single measure of concordance.

### Alignment-Expansion Framework

To facilitate interpretation of national adaptation patterns, countries were positioned on an Alignment-Expansion plane defined by their Alignment Ratio and Expansion Ratio. Countries were classified using the sample means of both measures as cut-points, generating four adaptation modes: restrictive/constrained adoption (low alignment, low expansion), normative dependence (high alignment, low expansion), autonomous divergence (low alignment, high expansion), and capacity-driven expansion (high alignment, high expansion). This framework provides a descriptive representation of how countries combine adoption of WHO-recommended medicines with expansion beyond the WHO benchmark.

### Health System Characteristics and National Adaptation of the WHO EML

We examined whether different country-level characteristics were associated with the two dimensions of adaptation.

Country-level explanatory variables were selected a priori to represent key structural domains that may influence national medicines selection. Variable selection was informed by literature on medicine policy and health systems: we considered population health needs, health system financing capacity, governance, and global integration (i.e. the extent to which a country is integrated into international economic, social, and political networks) (Bigdeli et al., 2013). These domains were represented by the under-five mortality rate (Mosley & Chen, 2003; World Bank, 2024), current health expenditure per capita (PPP) (World Health Organization, 2025), the Regulatory Quality Index (World Bank, 2024), and the Konjunkturforschungsstelle Globalisation Index (KOFGI) (Gygli et al., 2019; KOF Swiss Economic Institute), respectively.

WHO region and year of NEML publication were also included as explanatory variables. To approximate the health system and policy environment prevailing when medicine selection decisions were made, all country-level characteristics were measured two years before publication of each NEML.

### Statistical Analysis

Separate beta-regression models were respectively fitted for the Alignment Ratio and Expansion Ratio, which ranged between 0 and 1 without including boundary values (Ferrari & Cribari-Neto, 2004). Health expenditure per capita and under-five mortality were log-transformed to account for their right-skewed distributions.

Missing data were generally limited (<6% for each explanatory variable). However, we had to exclude Cook Islands, Niue, and the Democratic People’s Republic of Korea from regression analyses, as they had substantial missingness across key variables. Missing values in the remaining countries were addressed using multiple imputation by chained equations (MICE), using 20 imputed datasets (van Buuren, 2018). The imputation models included all variables in the primary analyses, the two outcomes, and six additional variables (child stunting prevalence, GDP per capita, urban population, population aged 65 years and above, internet use, and freedom of foreign movement (V-Dem)) to support the imputation. Estimates were combined using Rubin’s rules (Rubin, 1987).

Model diagnostics included assessment of multicollinearity using variance inflation factors and examination of residuals and influential observations using Cook’s distance. Sensitivity analyses assessed the robustness of the findings to alternative outcome definitions, medicine-matching approaches, temporal alignment of explanatory variables, and measures of national economic resources. The robustness of the Expansion Ratio was also assessed using absolute expansion counts and an offset-adjusted negative binomial model.

## Results

### Four-category framework of national adaptation

Classification according to the Alignment-Expansion framework identified four distinct patterns of adaptation. Fifty countries were categorized as restrictive/constrained adoption (low alignment, low expansion), thirty-five countries as normative dependence (high alignment, low expansion), sixteen countries as autonomous divergence (low alignment, high expansion) and thirty-six countries as capacity-driven expansion (high alignment, high expansion) shown in Figure 1. Alignment and Expansion Ratios varied substantially across countries, with mean values of 0.533 and 0.340, respectively. These findings indicate that adaptation of the WHO EML occurs through different combinations of adoption of WHO-recommended medicines and expansion beyond the WHO benchmark, rather than along a single continuum of concordance.

**Figure 1.**
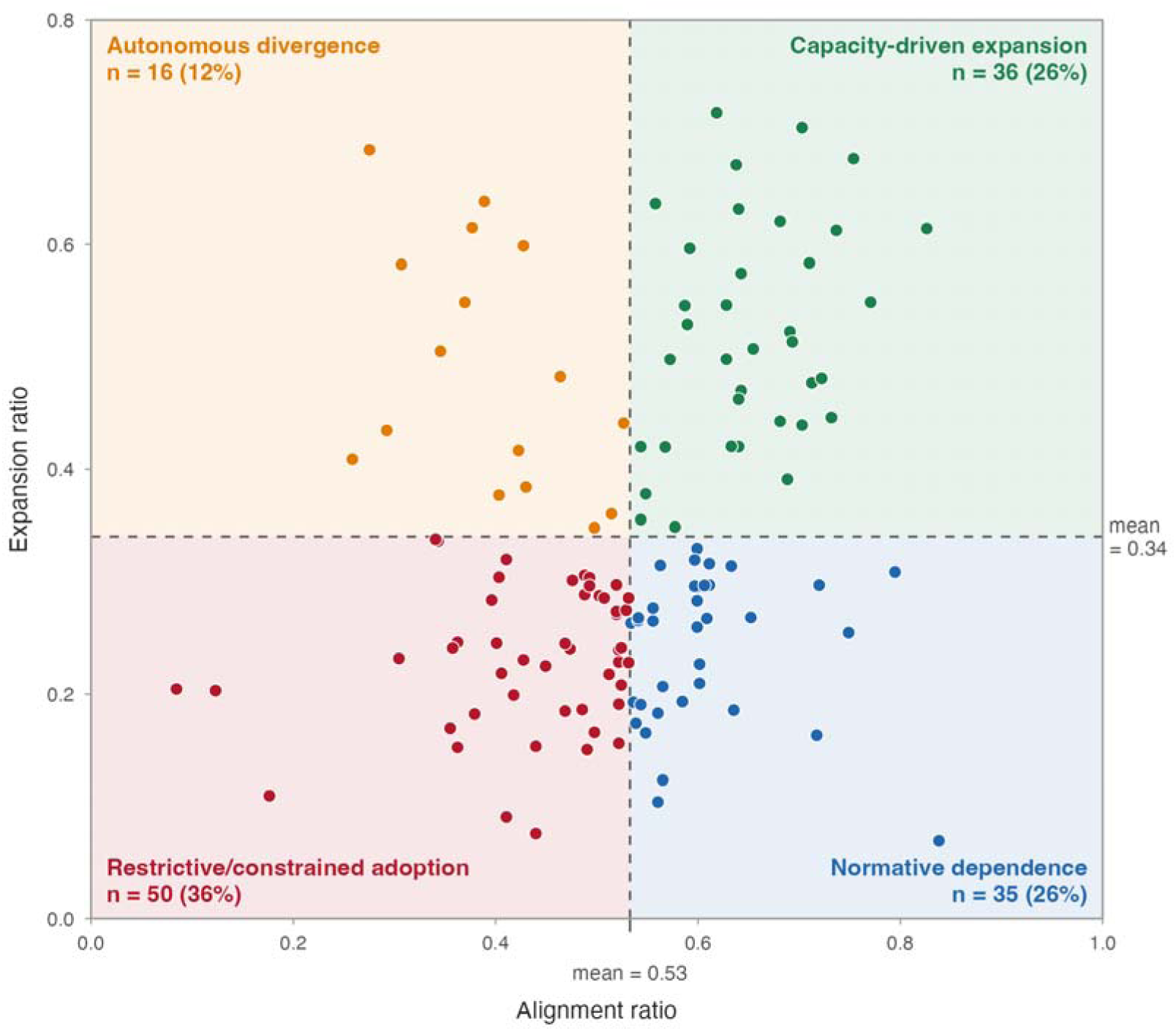
Alignment and expansion in national adaptation of the WHO Model List of Essential Medicines (WHO EML). Each point represents one NEML (n=137). Dashed lines indicate mean Alignment (0.533) and Expansion (0.340) Ratios used to define four descriptive adaptation modes. Alignment represents WHO EML medicines included nationally; expansion represents national medicines not included in the WHO EML: restrictive/constrained adoption (low alignment, low expansion; red), normative dependence (high alignment, low expansion; blue), autonomous divergence (low alignment, high expansion; yellow) and capacity-driven expansion (high alignment, high expansion; green). Countries may simultaneously exhibit high alignment and high expansion, indicating substantial adoption of WHO-recommended medicines alongside extensive additions reflecting national priorities. The typology is descriptive and does not imply a hierarchy of performance nor specific causal pathways underlying national medicines selection.

Restrictive/constrained adoption was most common in the African and Western Pacific Regions, whereas Capacity-driven expansion was more common in the Region of the Americas and Eastern Mediterranean Region. Countries in the European Region were primarily classified as Capacity-driven expansion or Autonomous divergence, while South-East Asia showed a relatively balanced distribution across the four categories (Figure 2).

**Figure 2.**
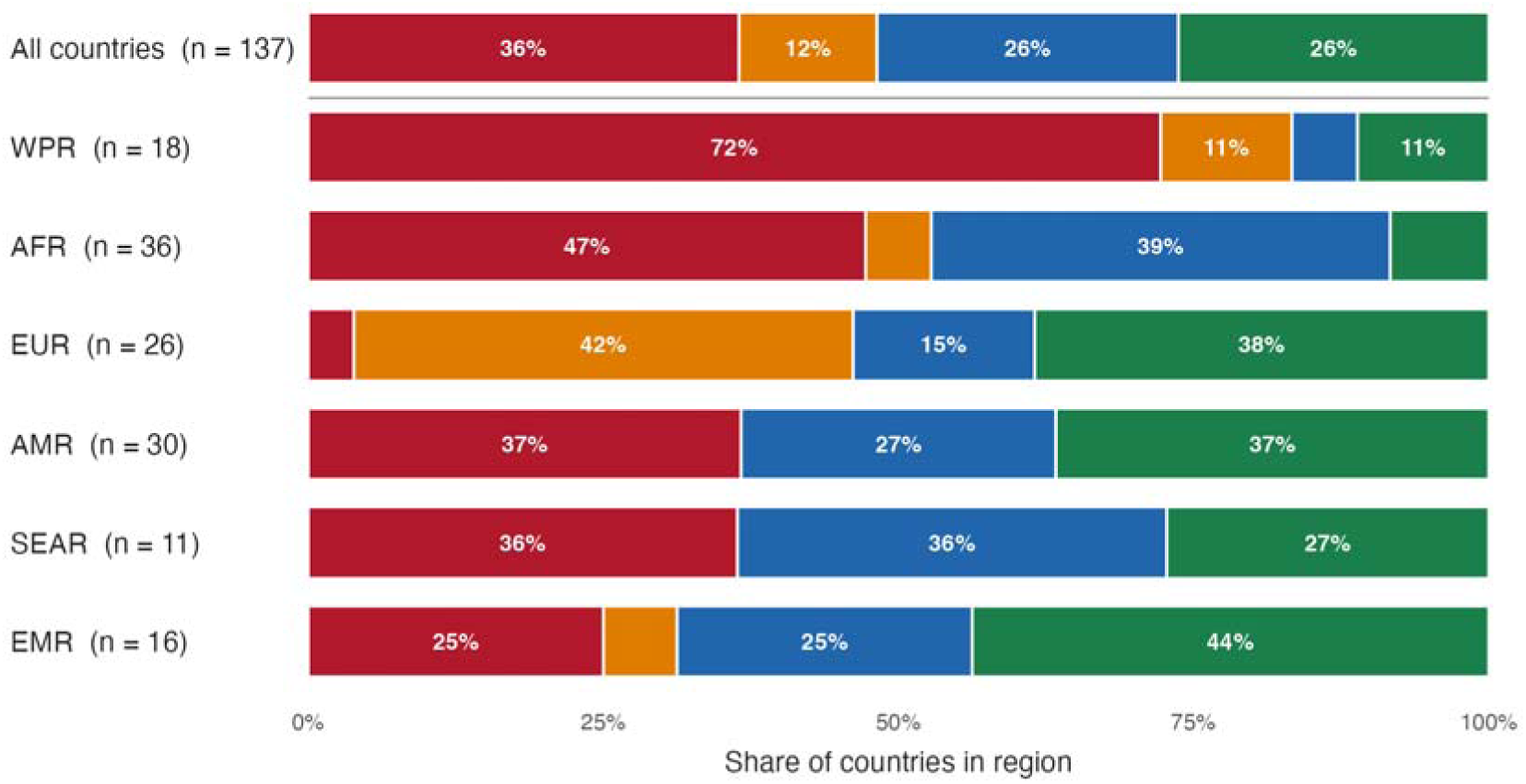
Distribution of national adaptation modes across WHO regions. Percentage of NEMLs (n=137) within each WHO region classified into the four adaptation modes. Classification used the mean Alignment Ratio (0.533) and Expansion Ratio (0.340) as thresholds. Values represent the proportion of countries within each region assigned to each mode. Regions differ markedly in the distribution of adaptation modes. Colours indicate restrictive/constrained adoption (red), normative dependence (blue), autonomous divergence (yellow), and capacity-driven expansion (green). Regional distributions illustrate differences in national adaptation patterns across WHO regions.

**Figure 3.**
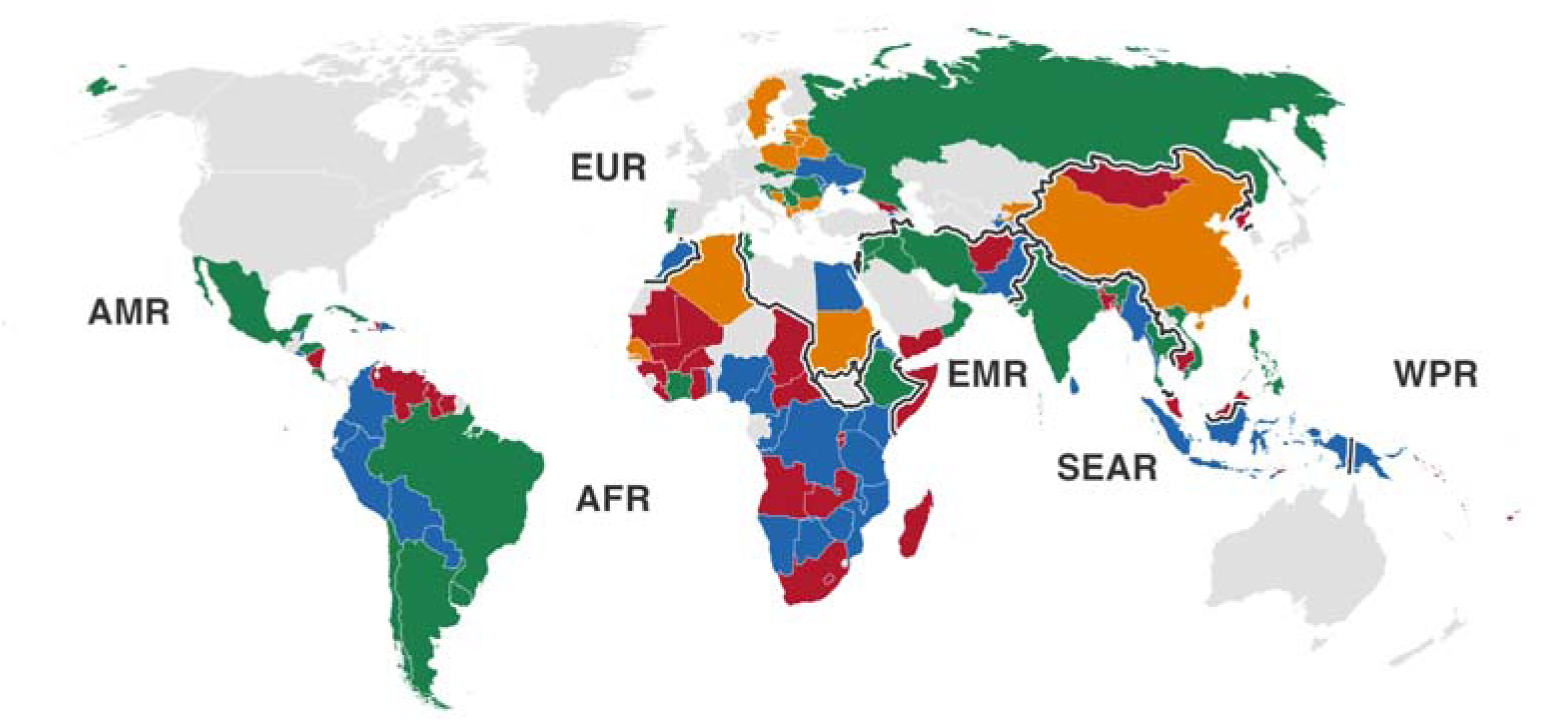
Geographic distribution of national EML adaptation modes. Geographic distribution of the four adaptation modes among 137 countries. Countries were classified according to alignment with the 2017 WHO EML and expansion beyond it, using the sample mean Alignment Ratio (0.533) and Expansion Ratio (0.340) as thresholds. Colours indicate restrictive/constrained adoption (red), normative dependence (blue), autonomous divergence (yellow), and capacity-driven expansion (green). Regional distributions illustrate differences in national adaptation patterns across WHO regions. Grey indicates countries for which no classification was available (no data). The geographic distribution indicates that national adaptation of the WHO EML follows diverse patterns that cannot be explained solely by regional affiliation, with neighbouring countries often displaying different approaches to balancing adherence to global recommendations and adaptation to national priorities.

### Factors associated with Alignment

Higher alignment was associated with lower under-five mortality (β = −0.294, 95% CI: −0.497 to −0.091, *p* = 0.005), greater globalization (β = 0.013, 95% CI: 0.000 to 0.026, *p* = 0.045), and more recent NEMLs were also associated with higher alignment (β = 0.046, 95% CI: 0.014 to 0.078, *p* = 0.005). Expenditure per capita showed borderline statistical significance (β = −0.158, CI: −0.315 to 0.000, *p* = 0.050). Overall, WHO region was significantly associated with alignment (pooled likelihood-ratio test, P = 0.0003). Compared with the African Region, countries in the European Region (β = −0.478, CI: -0.856 to –0.101) and Western Pacific Region (β = −0.406, CI: -0.726 to –0.086) had significantly lower alignment ratios, while no significant differences were observed for the other regions (Table 1).

**Table 1.** Beta regression analysis of country-level factors associated with Alignment with the WHO Model List of Essential Medicines;

| term |  | estimate | p.value | 95% confidence interval |
|---|---|---|---|---|
| Under-five mortality (log-transformed) |  | -0.294 | 0.005 | -0.497 to -0.091 |
| Health expenditure per capita, PPP (log-transformed) |  | -0.158 | 0.05 | -0.315 to 0.000 |
| KOF Globalisation Index |  | 0.013 | 0.045 | 0.000 to 0.026 |
| Regulatory quality |  | -0.137 | 0.096 | -0.298 to 0.025 |
| WHO region (reference: African Region) |  |  | Overall p value < 0.001 |  |
|  | Americas | 0.089 |  | -0.210 to 0.388 |
|  | Eastern Mediterranean | 0.14 |  | -0.170 to 0.450 |
|  | Europe | -0.478 |  | -0.856 to -0.101 |
|  | South-East Asia | -0.113 |  | -0.463 to 0.237 |
|  | Western Pacific | -0.406 |  | -0.726 to -0.086 |
| NEML publication year |  | 0.046 | 0.005 | 0.014 to 0.078 |

KOF, KOF Swiss Economic Institute; PPP, purchasing power parity; WHO, World Health Organization; NEML. National essential medicine list.

### Factors associated with Expansion

Higher expansion beyond the WHO EML was associated with lower under-five mortality (β = −0.481, 95% CI: −0.697 to −0.264, p < 0.001). No significant associations were observed between expansion and the other explanatory variables. Overall, WHO region was significantly associated with expansion (pooled likelihood-ratio test, P = 0.0024). Compared with the African Region, countries in the Region of the Americas (β = −0.619, 95% CI: −0.945 to −0.294), European Region (β = −0.470, 95% CI: −0.876 to −0.064), South-East Asia Region (β = −0.502, 95% CI: −0.895 to −0.109), and Western Pacific Region (β = −0.591, 95% CI: −0.942 to −0.239) had significantly lower expansion ratios. No significant difference was observed for the Eastern Mediterranean Region compared with the African Region (Table 2).

**Table 2.** Factors associated with expansion beyond the WHO Model List of Essential Medicines;

| **term** |  | **estimate** | **p.value** | **95% confidence interval** |
|---|---|---|---|---|
| Under-five mortality (log) |  | -0.481 | < 0.001 | -0.697 to -0.264 |
| Health expenditure per capita, PPP (log) |  | 0.140 | 0.10 | -0.026 to 0.305 |
| KOF Globalisation Index |  | 0.001 | 0.93 | -0.013 to 0.014 |
| Regulatory quality |  | -0.053 | 0.53 | -0.220 to 0.114 |
| WHO region (reference: African Region) |  |  | Overall p value : 0.002 |  |
|  | Americas | -0.619 |  | -0.945 to -0.294 |
|  | Eastern Mediterranean | -0.234 |  | -0.567 to 0.099 |
|  | Europe | -0.470 |  | -0.876 to -0.064 |
|  | South-East Asia | -0.502 |  | -0.895 to-0.109 |
|  | Western Pacific | -0.591 |  | -0.942 to -0.239 |
| NEML publication year |  | -0.015 | 0.40 | -0.049 to 0.020 |

KOF, KOF Swiss Economic Institute; PPP, purchasing power parity; WHO, World Health Organization; NEML. National essential medicine list.

Sensitivity analyses generally yielded results consistent with the primary findings, with some variation in statistical significance across specifications; the Expansion Ratio was also strongly correlated with absolute expansion count (Spearman’s ρ = 0.958), and 91% of countries retained the same adaptation classification when absolute expansion count was used instead.

## Discussion

The findings illustrate a limitation of viewing national implementation of the WHO Model List as a one-dimensional construct. Similar levels of similarity to the WHO benchmark may conceal qualitatively different adaptation strategies. Some countries primarily diverge through incomplete adoption of WHO-recommended medicines, whereas others diverge through substantial additions beyond the WHO EML, and many through a combination of both. By separating alignment from expansion, the analysis provides a more informative description of national adaptation and highlights that differences between national lists and the WHO EML should not automatically be interpreted as reflecting the same underlying policy processes. Understanding these processes may become increasingly important as the WHO Model List continues to evolve, while countries simultaneously face changing economic constraints, epidemiological transitions, and evolving priorities for medicine selection and financing.

### Factors associated with alignment and expansion

The association between under-five mortality and lower alignment may indicate that translating a global normative standard into national policy depends partly on health-system capacity, rather than epidemiological need alone. The positive association with globalization may reflect greater opportunities for policy learning and diffusion of international recommendations through global governance networks during a period characterized by increasing global interconnectedness (Dolowitz & Marsh, 2000). Regional patterns were also observed, although these should be interpreted cautiously. Rather than suggesting an independent effect of region itself, they may reflect shared historical, economic, and policy trajectories among countries within regions, while national adaptation ultimately remains shaped by country-specific circumstances and priorities (Saxena et al., 2026; Steiner et al., 2020; Taglione & Persaud, 2021). The borderline association with health expenditure may suggest that greater resources do not necessarily translate into closer alignment, as countries may instead use HTA, clinical guidelines, and historic reimbursement trends to select medicines beyond the WHO EML (Glassman, 2017).

In contrast, expansion showed relatively few consistent associations with country-level characteristics. This finding suggests that additions beyond the WHO EML may be less strongly determined by health-system resources or development indicators than by national priority-setting processes. Expansion is likely influenced by factors such as historical formulary development, professional preferences, reimbursement policies, and other contextual considerations that are not readily captured by broad ecological indicators (Glassman et al., 2016; Peacocke et al., 2022). Countries facing greater constraints may prioritize securing access to the WHO baseline before allocating resources to additional medicines.

### Strengths and limitations

Several limitations should be noted. First, the analysis used the 2017 WHO EML and corresponding NEMLs. This allowed us to apply the proposed framework to a database that has been used in several previous comparative analyses and to explore whether alignment and expansion may provide useful descriptors for interpreting NEMLs in future monitoring exercises. Second, the cross-sectional design and limited set of country-level covariates preclude causal interpretation and may leave relevant determinants unmeasured. Third, the dataset did not account for therapeutic alternatives. Medicines included in national lists that serve as substitutes for WHO-recommended medicines were treated as distinct medicines rather than therapeutic equivalents. This may have resulted in an underestimation of alignment and a corresponding overestimation of expansion. Finally, medicine selection does not necessarily reflect availability or health outcomes.

## Conclusion

The framework shifts attention from whether countries conform to the WHO EML toward how they adapt it to national circumstances, providing a non-normative lens to distinguish incomplete adoption from deliberate expansion and potentially facilitating policy dialogue on national medicines selection. More broadly, it suggests a move away from interpreting differences between national essential medicines lists and the WHO EML as a simple measure of divergence. This perspective may be particularly relevant as NEMLs increasingly serve not only as policy tools but also as indicators for monitoring progress on essential medicines. In the WHO GPW-14 results framework, indicators related to national essential medicines lists contribute to monitoring Member State progress on access to essential medicines, underscoring the importance of understanding what changes in these lists represent in practice (World Health Organization, 2025). Future research should examine whether adaptation patterns are associated with medicine availability, health-system performance, health outcomes, and equitable access, since NEML inclusion does not guarantee access.

## Author contributions

Yukari Kohno: Conceptualization, Methodology, Data curation, Formal analysis, Visualization, Writing – original draft.

Janne Anton Markus Estill: Methodology, Writing – review & editing.

Lorenzo Moja: Conceptualization, Writing – review & editing.

Olivia Keiser: Supervision, Methodology, Writing – review & editing.

## Disclosure statement

No potential conflict of interest was reported by the authors.

## Funding details

This research received no specific funding.

## Declaration of generative AI use

ChatGPT (OpenAI; o3) was used during manuscript preparation to support brainstorming on statistical approaches and the exploration of possible interpretations of the findings, as well as language editing and refinement. Claude Code (Anthropic; Claude Opus 4.8) was used to assist in writing the statistical analysis code. The authors independently reviewed and verified the code and all statistical analyses, and determined the final interpretation and conclusions. All AI-assisted content was reviewed and verified by the authors, including the accuracy and appropriateness of references. The authors confirm the originality and accuracy of the manuscript, have reviewed the applicable terms of use of the AI tools, and take full responsibility for the integrity of the entire manuscript.

## Data availability statement

The medicine-level data used in this study are available from the Global Essential Medicines Database, originally published by Persaud et al. (2019) and available at https://figshare.com/articles/dataset/GlobalEssentialMedicinesDatabase_xlsx/7814246. The WHO Model List of Essential Medicines 2017 was used as the reference benchmark. Country-level explanatory variables were compiled from publicly available sources, as detailed in the Methods section. The analytical dataset used in this study was derived by integrating these data sources and constructing the study-specific outcome measures.

## Ethics approval

Ethics approval was not required for this study because it involved secondary analysis of publicly available, country-level data and did not involve human participants, identifiable personal data, or animals.

## Consent to participate

Consent to participate was not applicable because this study involved secondary analysis of publicly available, country-level data and did not involve human participants.

## Notes

### Competing Interest Statement

The authors have declared no competing interest.

## References

Bigdeli, M., Jacobs, B., Tomson, G., Laing, R., Ghaffar, A., Dujardin, B., & Van Damme, W. (2013). Access to medicines from a health system perspective. Health Policy and Planning, 28(7), 692–704. 10.1093/heapol/czs108

Dolowitz, D. P., & Marsh, D. (2000). Learning from Abroad: The Role of Policy Transfer in Contemporary Policy-Making. Governance, 13(1), 5–23. 10.1111/0952-1895.00121

Ferrari, S., & Cribari-Neto, F. (2004). Beta Regression for Modelling Rates and Proportions. Journal of Applied Statistics, 31(7), 799–815. 10.1080/0266476042000214501

Glassman, A. (2017). What’s in, What’s Out: Designing Benefits for Universal Health Coverage (1st ed.). Center for Global Development.

Glassman, A., Giedion, U., Sakuma, Y., & Smith, P. C. (2016). Defining a Health Benefits Package: What Are the Necessary Processes? Health Systems & Reform, 2(1), 39–50. 10.1080/23288604.2016.1124171

Gygli, S., Haelg, F., Potrafke, N., & Sturm, J.-E. (2019). The KOF Globalisation Index – revisited. The Review of International Organizations, 14(3), 543–574. 10.1007/s11558-019-09344-2

Heredia, C., Ul Haq, M. Z., Cappello, B., Malik, F., Penazzato, M., Moja, L., & Persaud, N. (2025). Pediatric formulations in national essential medicines lists: a cross-sectional study. Front Pediatr, 13, 1566841. 10.3389/fped.2025.1566841

Jenei, K. (2025). The Political Economy of the World Health Organization Model Lists of Essential Medicines. Milbank Quarterly, 103(1), 52–99. 10.1111/1468-0009.70001

KOF Swiss Economic Institute. KOF Globalisation Index 2023 (Version 2023) [Data set]. ETH Zurich. Available at: https://kof.ethz.ch/en/forecasts-and-indicators/indicators/kof-globalisation-index.html [Accessed: October 29,2026].

Laing, R., Waning, B., Gray, A., Ford, N., & Hoen, E. (2003). 25 years of the WHO essential medicines lists: progress and challenges. The Lancet, 361(9370), 1723–1729. 10.1016/S0140-6736(03)13375-2

Mosley, W. H., & Chen, L. C. (2003). An analytical framework for the study of child survival in developing countries. Bulletin of the World Health Organization, 81(2), 140–145. https://www.ncbi.nlm.nih.gov/pubmed/12756980

Peacocke, E. F., Myhre, S. L., Foss, H. S., & Gopinathan, U. (2022). National adaptation and implementation of WHO Model List of Essential Medicines: A qualitative evidence synthesis. PLoS Medicine, 19(3), e1003944. 10.1371/journal.pmed.1003944

Persaud, N., Jiang, M., Shaikh, R., Bali, A., Oronsaye, E., Woods, H., Drozdzal, G., Rajakulasingam, Y., Maraj, D., Wadhawan, S., Umali, N., Wang, R., McCall, M., Aronson, J. K., Pluddemann, A., Moja, L., Magrini, N., & Heneghan, C. (2019). Comparison of essential medicines lists in 137 countries. Bulletin of the World Health Organization, 97(6). 10.2471/BLT.18.222448

Piggott, T., Nowak, A., Brignardello-Petersen, R., Cooke, G. S., Huttner, B., Schünemann, H. J., Persaud, N., Magrini, N., & Moja, L. (2022). Global status of essential medicine selection: a systematic comparison of national essential medicine lists with recommendations by WHO. BMJ Open, 12(2), e053349. 10.1136/bmjopen-2021-053349

Rubin, D. B. (1987). *Multiple Imputation for Nonresponse in Surveys* (1 ed.). Wiley. 10.1002/9780470316696

Saxena, E., Kohler, J. C., Thorpe, K. E., & Persaud, N. (2026). Assessing the national essential medicines list selection processes: Instrument development and testing. PloS One, 21(2), e0342750. 10.1371/journal.pone.0342750

Steiner, L., Maraj, D., Woods, H., Jarvis, J., Yaphe, H., Adekoya, I., Bali, A., & Persaud, N. (2020). A comparison of national essential medicines lists in the Americas. Revista Panamericana de Salud Publica, 44, e5. 10.26633/RPSP.2020.5

Taglione, M. S., & Persaud, N. (2021). Assessing variation among the national essential medicines lists of 21 high-income countries: a cross-sectional study. BMJ Open, 11(8), e045262. 10.1136/bmjopen-2020-045262

van Buuren, S. (2018). Flexible Imputation of Missing Data (2nd ed.). Chapman & Hall/CRC. https://stefvanbuuren.name/fimd/

W Wirtz, V. J., Hogerzeil, H. V., Gray, A. L., Bigdeli, M., de Joncheere, C. P., Ewen, M. A., Gyansa-Lutterodt, M., Jing, S., Luiza, V. L., Mbindyo, R. M., Möller, H., Moucheraud, C., Pécoul, B., Rägo, L., Rashidian, A., Ross-Degnan, D., Stephens, P. N., Teerawattananon, Y., ’t Hoen, E. F. M., . . . Reich, M. R. (2017). Essential medicines for universal health coverage. Lancet, 389(10067), 403–476. 10.1016/S0140-6736(16)31599-9

World Bank. (2024). Worldwide Governance Indicators [database on the Internet]. Washington (DC): World Bank. Available at: https://www.worldbank.org/en/publication/worldwide-governance-indicators [Accessed: December 2, 2024]. https://www.worldbank.org/en/publication/worldwide-governance-indicators

World Bank. (2024). World Development Indicators [database on the Internet]. Washington (DC): World Bank. Available at: https://databank.worldbank.org/source/world-development-indicators. [Accessed: December 2, 2024].

World Health Organization (2025). Global Health Expenditure Database [database on the Internet]. Geneva: World Health Organization. Available at: https://apps.who.int/nha/database/Select/Indicators/en [Accessed: May 26, 2026].

World Health Organization. (2017). WHO Model List of Essential Medicines, 20th List (April 2017). World Health Organization. https://www.who.int/publications/i/item/eml-20 *[Accessed: October 29, 2026]*.

World Health Organization. (2026). Anatomical Therapeutic Chemical (ATC) Classification. World Health Organization*. Available at:* https://www.who.int/tools/atc-ddd-toolkit/atc-classification *[Accessed: October 29, 2026]*

World Health Organization. Selection of essential medicines at country level: using the WHO Model List of Essential Medicines to update a national essential medicines list. Geneva: World Health Organization; 2020. *Licence: CC BY-NC-SA 3.0 IGO. Available at:* https://iris.who.int/server/api/core/bitstreams/76b3d5fc-5920-458a-9f4b-b38565b088e0/content *[Accessed: October 29, 2026]*

World Health Organization. World Health Organization. A global health strategy for 2025–2028: advancing equity and resilience in a turbulent world. Fourteenth General Programme of Work (GPW 14). Geneva: World Health Organization; 2025. *Available at:* https://www.who.int/publications/i/item/9789240101012 *[Accessed: October 29, 2026]*.

